# Real World Evidence of Neutralizing Monoclonal Antibodies for Preventing Hospitalization and Mortality in COVID-19 Outpatients

**DOI:** 10.1101/2022.01.09.22268963

**Authors:** Matthew K. Wynia, Laurel E. Beaty, Tellen D. Bennett, Nichole E. Carlson, Christopher B. Davis, Bethany M. Kwan, David A. Mayer, Toan C. Ong, Seth Russell, Jeffrey Steele, Heather R. Stocker, Adane F. Wogu, Richard D. Zane, Ronald J. Sokol, Adit A. Ginde

## Abstract

**Background:** Neutralizing monoclonal antibodies (mAbs) are authorized for early symptomatic COVID-19 patients. Whether mAbs are effective against the SARS-CoV-2 Delta variant, among vaccinated patients, or for prevention of mortality remains unknown.

**Objective:** To evaluate the effectiveness of mAb treatment in preventing progression to severe disease during the Delta phase of the pandemic and based on key baseline risk factors.

**Design, Setting, and Patients:** Observational cohort study of non-hospitalized adult patients with SARS-CoV-2 infection from November 2020-October 2021, using electronic health records from a statewide health system plus state-level vaccine and mortality data. Using propensity matching, we selected approximately 2.5 patients not receiving mAbs for each patient who received mAbs.

**Exposure:** Neutralizing mAb treatment under emergency use authorization

**Main Outcomes:** The primary outcome was 28-day hospitalization; secondary outcomes included mortality and severity of hospitalization.

**Results:** Of 36,077 patients with SARS-CoV-2 infection, 2,675 receiving mAbs were matched to 6,677 not receiving mAbs. Compared to mAb-untreated patients, mAb-treated patients had lower all-cause hospitalization (4.0% vs 7.7%; adjusted OR 0.48, 95%CI 0.38-0.60) and all-cause mortality (0.1% vs. 0.9%; adjusted OR 0.11, 95%CI 0.03-0.29) to day 28; differences persisted to day 90. Among hospitalized patients, mAb-treated patients had shorter hospital length of stay (5.8 vs. 8.5 days) and lower risk of mechanical ventilation (4.6% vs. 16.6%). Relative effectiveness was similar in preventing hospitalizations during the Delta variant phase (adjusted OR 0.35, 95%CI 0.25-0.50) and across subgroups. Lower number-needed-to-treat (NNT) to prevent hospitalization were observed for subgroups with higher baseline risk of hospitalization (e.g., multiple comorbidities (NNT=17) and not fully vaccinated (NNT=24) vs. no comorbidities (NNT=88) and fully vaccinated (NNT=81).

**Conclusion:** Real-world evidence demonstrated mAb effectiveness in reducing hospitalization among COVID-19 outpatients, including during the Delta variant phase, and conferred an overall 89% reduction in 28-day mortality. Early outpatient treatment with mAbs should be prioritized, especially for individuals with highest risk for hospitalization.

## INTRODUCTION

High rates of coronavirus disease 2019 (COVID-19) transmission and illness persist, especially among unvaccinated individuals, as well as those with waning vaccine or infection-related immunity, such as older adults or those with certain chronic medical conditions.^1,2^ Neutralizing monoclonal antibody (mAb) treatment provides immediate passive immunity against severe acute respiratory syndrome coronavirus-2 (SARS-CoV-2), the virus that causes COVID-19.

Several mAb products have received emergency use authorization (EUA) from the US Food and Drug Administration.^3^ These authorizations were based on early Phase II/III randomized controlled trials that demonstrated reduction in a combined endpoint of hospitalization or death among high-risk outpatients with early symptomatic infection, though these trials were small in size with few deaths and conducted before the emergence of the Delta variant or widespread availability of vaccines against SARS-CoV-2.^4-6^

Once a promising therapeutic agent has been authorized for emergency use, it becomes more challenging to recruit patients into randomized controlled trials, as patients may seek active therapy and clinicians may view randomization to placebo as unethical.^7^ Consequently, studies of mAbs following EUA have primarily been small observational trials, confirming reduced hospitalization rates but not large enough to detect a mortality benefit nor to assess any potential heterogeneity of mAb treatment effects by comorbid conditions or vaccination status.^8-10^ The latter information could be especially useful in policymaking about how best to allocate limited access to mAb treatment during shortages.^11,12^ Furthermore, no published studies have yet directly evaluated the effectiveness of currently available mAbs against the Delta variant of SARS-CoV-2, which arose in summer 2021 in the US.

The rapidly-evolving nature of the COVID-19 pandemic, including both the emergence of new variants of the virus and use of EUAs allowing early access to novel therapeutics, makes it critical to build robust research platforms for real-world evidence generation.^13,14^ In early 2021, we created a real-world evidence platform to assess the ongoing clinical impacts of mAb therapy on high-risk outpatients with early symptomatic COVID-19.

Our study objective was to evaluate the effectiveness of mAb treatment and progression to severe disease, including hospitalization, severity of hospitalization, and mortality. The goal of the overall platform was to include changes in the pandemic, including emergence of new variants, in near real-time with sufficient power to assess potential mortality benefits and effectiveness among patients with various risk factors for progression to severe disease, including vaccination status.

## METHODS

### Study Oversight and Data Sources

We conducted a propensity-matched observational cohort study, as part of a statewide implementation/effectiveness pragmatic trial, in a collaboration between University of Colorado researchers, University of Colorado Health (UCHealth) leaders, and the Colorado Department of Public Health and Environment (CDPHE). The study was approved by the Colorado Multiple Institutional Review Board with a waiver of informed consent. We obtained data from the electronic health record (EHR; Epic, Verona, WI) of UCHealth, the largest health system in Colorado with 13 hospitals around the state and 141,000 annual hospital admissions. EHR data were merged with statewide data on vaccination status from the Colorado Comprehensive Immunization Information System and mortality from Colorado Vital Records.

### Patient Population Studied

We included patients diagnosed with SARS-CoV-2 infection between November 20, 2020 and October 7, 2021 allowing for at least 28 days of follow-up as of November 4, 2021 (n=36,077), identified using EHR-based date of SARS-CoV-2 positive testing (by polymerase chain reaction or antigen tests) or date of administration of mAb treatment (if no SARS-CoV-2 test result date available). The decision to seek mAb treatment was made by patients and clinicians, and a state-wide referral system was established by CDPHE to facilitate patient referrals to facilities for mAb infusion.^15^ We did not exclude patients solely for lack of EUA eligibility based on EHR data, because not all eligibility criteria were consistently available in the EHR (see additional Methods in the Supplement). We excluded patients who received mAb treatment on the same day of or during hospitalization, as these patients already had the primary outcome. Logistic regression was used for propensity score estimation^16^ with nearest neighbor matching^17^ applied to select an approximate 2.5:1 mAb-untreated to mAb-treated matched cohort. Matching factors included baseline demographics, clinical variables, and time (see additional Methods in the Supplement). The primary analysis cohort included patients with a documented mAb administration date (n=2,675) and propensity-matched controls who did not receive mAb treatment (n=6,677). We assessed effectiveness of matching using standardized mean differences.^18^

### Outcomes

The primary outcome was all-cause hospitalization within 28 days of a positive SARS-CoV-2 test, obtained from EHR data. Secondary outcomes included all-cause hospitalization to day 90, all-cause mortality to days 28 and 90, and emergency department (ED) visits to day 28. Among those hospitalized, outcomes included disease severity based on maximum level of respiratory support, hospital and intensive care unit (ICU) length of stay (LOS), and rates of ICU admission, mechanical ventilation, and in-hospital mortality. Subgroups examined for the primary outcome included age, sex, combined race/ethnicity, insurance status, immunocompromised status, total number of other comorbidities, specific comorbidities, vaccination status, pandemic phase, and type of mAb treatment.

### Variable Definitions

The treatment variable was mAb administration and the primary starting point (time zero) was the date of any SARS-CoV-2 positive test. We imputed missing test dates based on the distribution of observed mAb administration dates (see additional Methods in the Supplement). Hospitalization was defined as any inpatient or observation encounter documented in the EHR. ED visits were defined as any visit to the ED, with or without an associated inpatient or observation encounter. Presence of comorbid conditions were determined using a 90-day look back period in the EHR using established algorithms and immunosuppressed status was further validated by manual chart reviews (see additional Methods in the Supplement). COVID-19 disease severity was estimated using ordinal categories of respiratory support requirements at an encounter level, based on the highest level of support received among the following types (in increasing order): no oxygen, standard (nasal cannula/face mask) oxygen, high-flow nasal cannula or non-invasive ventilation, and invasive mechanical ventilation.^19^ In-hospital mortality was the highest level of disease severity.

Pandemic phase was categorized by SARS-CoV-2 positive date based on the prevalent variant in Colorado as Pre-Alpha (November 2020 - February 2021), Alpha (March 2021 – June 2021), and Delta (July 2021 – December 2021). No virus sequencing results were available on an individual patient basis. Vaccination status at the time of SARS-CoV-2 positive date was categorized as fully vaccinated (at least 14 days after primary vaccine series) or not fully vaccinated, which included partially vaccinated (receipt of at least vaccine dose but completed primary series) or not known to be vaccinated. MAb treatments included bamlanivimab (Eli Lilly), casirivimab + imdevimab (Regeneron), bamlanivimab + etesevimab (Eli Lilly), and sotrovimab (GlaxoSmithKline) (see additional Methods in the Supplement for more details).

### Statistical analysis

We present results descriptively and adjusted for potential confounders. All regression models for outcomes were adjusted for age, sex, race/ethnicity, insurance status, BMI, immunocompromised status, number of comorbidities, pandemic phase, and vaccination status. For binary outcomes such as hospitalization, we used logistic regression to determine odds of the outcome. For count outcomes such as LOS, we used Poisson regression to estimate incidence rates. We analyzed disease severity using ordinal logistic regression to estimate the proportional odds. We constructed cumulative incidence curves using Kaplan-Meier estimates to visually assess temporal trends by treatment status.

We conducted subgroup analyses to estimate heterogeneity of treatment effect for the primary outcome of all-cause hospitalization to day 28. For each subgroup, we calculated unadjusted rates of hospitalization, number needed to treat (NNT) to prevent one hospitalization (based on absolute risk reduction in unadjusted hospitalization rates), and adjusted relative odds of hospitalization. Results are presented as effect sizes, with 95% confidence intervals, and were not adjusted for multiple comparisons.

Two sensitivity analyses were performed. The first included only EUA-eligible subjects as verified by available EHR data. The second used a more conservative imputation method for missing SARS-CoV-2 positive test dates by assuming all missing positive test dates were ten days prior to the mAb administration date (the maximum time difference allowed by the EUA). All outcome models were repeated for these two cohorts and results compared with primary analyses. All statistical analyses were performed using R Statistical Software (version 3.6.0; R Foundation for Statistical Computing, Vienna, Austria).^20^

## RESULTS

### Characteristics of mAb-Treated and mAb-Untreated Cohorts

Of 36,077 patients with SARS-CoV-2 infection, 2,675 receiving mAbs were matched to 6,677 patients not receiving mAbs (**Appendix Figure 1** in the Supplement). The characteristics of mAb-treated and mAb-untreated patients in the primary cohort are presented in **Table 1**. The mAb-treated cohort generally reflects EUA criteria for use of mAbs, with many being older (40.7% were age ≥ 65 years), having higher BMI (50.1% with BMI over 25 kg/m^2^) and/or having one or more comorbidities (73.6%). While there were clinically important differences between mAb-treated and mAb-untreated patents in the full cohort (**Appendix Table 1** in the Supplement), propensity matching eliminated clinically meaningful differences between groups on matching variables (**Table 1, Appendix Table 2** in the Supplement). The mean time from positive SARS-CoV-2 test to receipt of mAb treatment was 3.7 days (SD 2.5).

**Table 1.** Baseline Characteristics by Monoclonal Antibody Treatment Status for Primary Matched Cohort.

| Characteristic | mAb-Treated<br>n=2675 | mAb-Untreated<br>n=6677 |
| --- | --- | --- |
| Age in years* |  |  |
| 18-54 years | 1018 (38.1%) | 3025 (45.3%) |
| 55-64 years | 569 (21.3%) | 1635 (24.5%) |
| ≥65 years | 1088 (40.7%) | 2017 (30.2%) |
| Female Sex* | 1453 (54.3%) | 3705 (55.5%) |
| Race/Ethnicity* |  |  |
| Non-Hispanic White | 2215 (82.8%) | 5323 (79.7%) |
| Hispanic | 264 (9.9%) | 775 (11.6%) |
| Non-Hispanic Black | 64 (2.4%) | 189 (2.8%) |
| Other | 132 (4.9%) | 390 (5.8%) |
| Insurance Status* |  |  |
| Private/Commercial | 1355 (50.7%) | 3840 (57.5%) |
| Medicare | 1052 (39.3%) | 1989 (29.8%) |
| Medicaid | 164 (6.1%) | 543 (8.1%) |
| None/Uninsured | 44 (1.6%) | 118 (1.8%) |
| Other/Unknown | 60 (2.2%) | 187 (2.8%) |
| Body mass index in kg/m<sup>2</sup>* |  |  |
| <18.5 | 23 (0.9%) | 60 (0.9%) |
| 18.5-24.9 | 362 (13.5%) | 875 (13.1%) |
| 25.0-29.9 | 571 (21.3%) | 1374 (20.6%) |
| ≥30.0 | 770 (28.8%) | 2013 (30.1%) |
| Missing | 949 (35.5%) | 2355 (35.3%) |
| Immunocompromised* | 809 (30.2%) | 1677 (25.1%) |
| Number of Other Comorbid Conditions* |  |  |
| 0 | 708 (26.5%) | 1837 (27.5%) |
| 1 | 681 (25.5%) | 1967 (29.5%) |
| ≥2 | 1286 (48.1%) | 2873 (43.0%) |
| Diabetes | 561 (21.0%) | 1173 (17.6%) |
| Cardiovascular Disease | 557 (20.8%) | 1290 (19.3%) |
| Pulmonary Disease | 891 (33.3%) | 2109 (31.6%) |
| Renal Disease | 344 (12.9%) | 607 (9.1%) |
| Hypertension | 1293 (48.3%) | 2881 (43.1%) |
| Obesity | 808 (30.2%) | 2073 (31.0%) |
| Vaccination Status |  |  |
| Not known to be vaccinated | 1620 (60.6%) | 4394 (65.8%) |
| Partially vaccinated | 148 (5.5%) | 485 (7.3%) |
| Fully vaccinated | 907 (33.9%) | 1798 (26.9%) |
| Pandemic Phase |  |  |
| Pre-alpha: Nov 2020 - Feb 2021 | 388 (14.5%) | 984 (14.7%) |
| Alpha: March 2021 - June 2021 | 615 (23.0%) | 1794 (26.9%) |
| Delta: July 2021 - Sep 2021 | 1672 (62.5%) | 3899 (58.4%) |
| Type of monoclonal antibody |  |  |
| Bamlanivimab | 413 (15.4%) | -- |
| Bamlanivimab + etesevimab | 87 (3.3%) | -- |
| Casirivimab + imdevimab | 2157 (80.6%) | -- |
| Sotrovimab | 18 (0.7%) | -- |

\* Variables used in the propensity matching. Abbreviations: mAb, monoclonal antibody

**Table 2.** Primary and Secondary Outcomes by Monoclonal Antibody Treatment Status.

| **Outcome** | **mAb-Treated** | **mAb-Untreated** | **Adjusted OR** | **95% CI** |
| --- | --- | --- | --- | --- |
| **Overall Sample Size** | **n=2675** | **n=6677** |  |  |
| All-Cause Hospitalization |  |  |  |  |
| 28-day (primary outcome) | 108 (4.0%) | 511 (7.7%) | 0.48 | (0.38, 0.60) |
| 90-day | 138 (5.2%) | 590 (8.8%) | 0.53 | (0.44, 0.65) |
| All-Cause Mortality |  |  |  |  |
| 28-day | 3 (0.1%) | 63 (0.9%) | 0.11 | (0.03, 0.29) |
| 90-day | 6 (0.2%) | 84 (1.3%) | 0.17 | (0.06, 0.35) |
| Any ED Visit to Day 28 | 501 (18.7%) | 1128 (16.9%) | 1.24 | (1.09, 1.40) |
| ED Visit leading to Hospitalization | 80/501 (16.0%) | 424/1128 (37.6%) | 0.29 | (0.21, 0.38) |
| **Hospitalized Sample Size** | **n=108** | **n=511** |  |  |
| Hospital LOS in days, mean (SD)* | 5.8 (6.5) | 8.5 (9.8) | 0.64 | (0.51, 0.82) |
| IMV or Death | 5 (4.6%) | 85 (16.6%) | 0.22 | (0.07, 0.52) |
| ICU Admission | 13 (12.0%) | 100 (19.6%) | 0.52 | (0.26, 0.97) |
| ICU LOS (days), mean (SD)* | 3.5 (2.8) | 8.6 (9.9) | 0.22 | (0.10, 0.48) |

\* Poisson regressions presented as adjusted incidence rate ratios with 95% confidence intervals
All regression models adjusted for age, sex, race/ethnicity, BMI, immunocompromised status, number of other comorbidities, insurance status, pandemic phase, and vaccination status
Abbreviations: mAb, monoclonal antibody; OR, odds ratio; CI, confidence interval; LOS, length of stay; ICU, intensive care unit; SD, standard deviation

**Figure 1.**
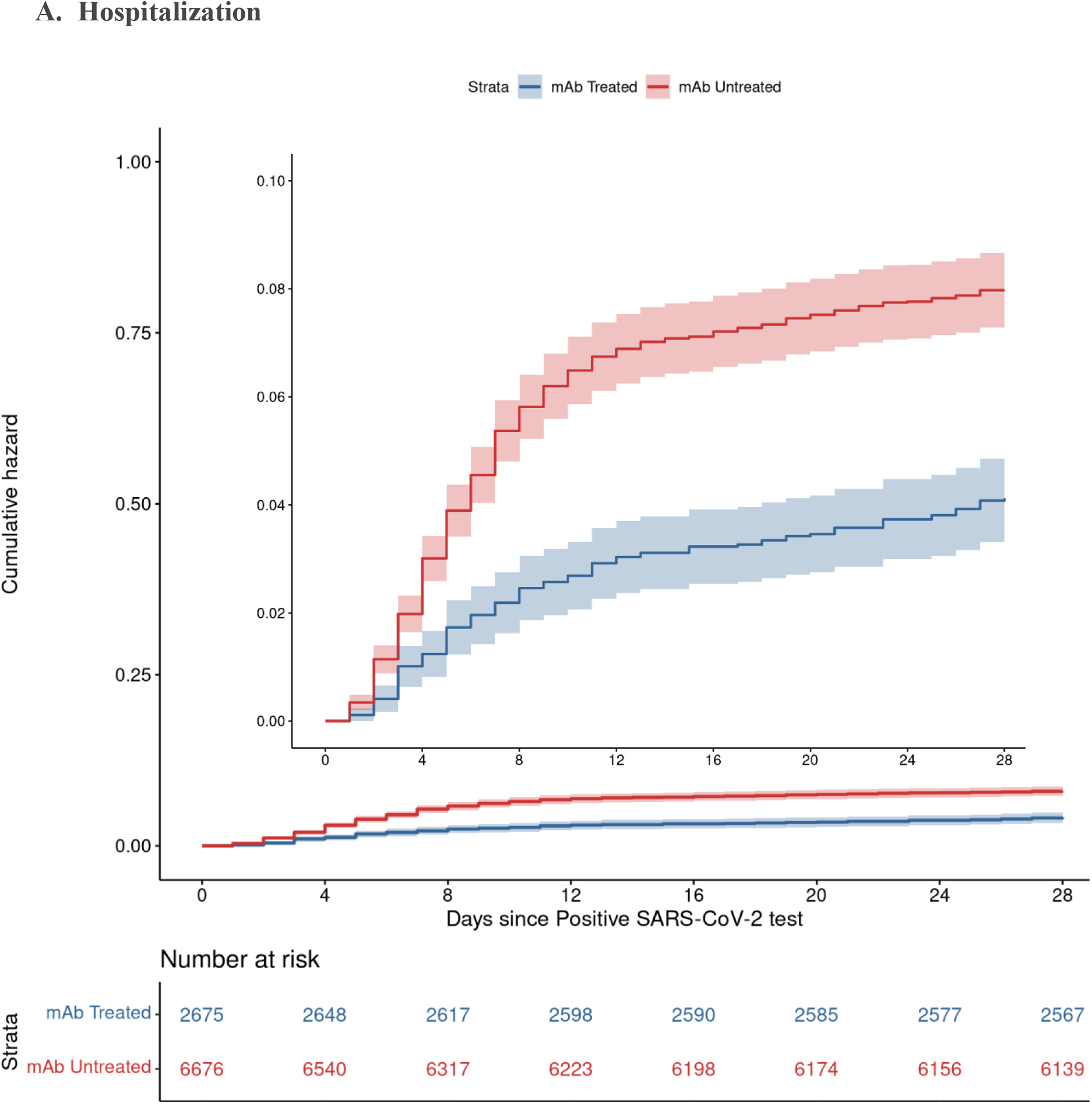
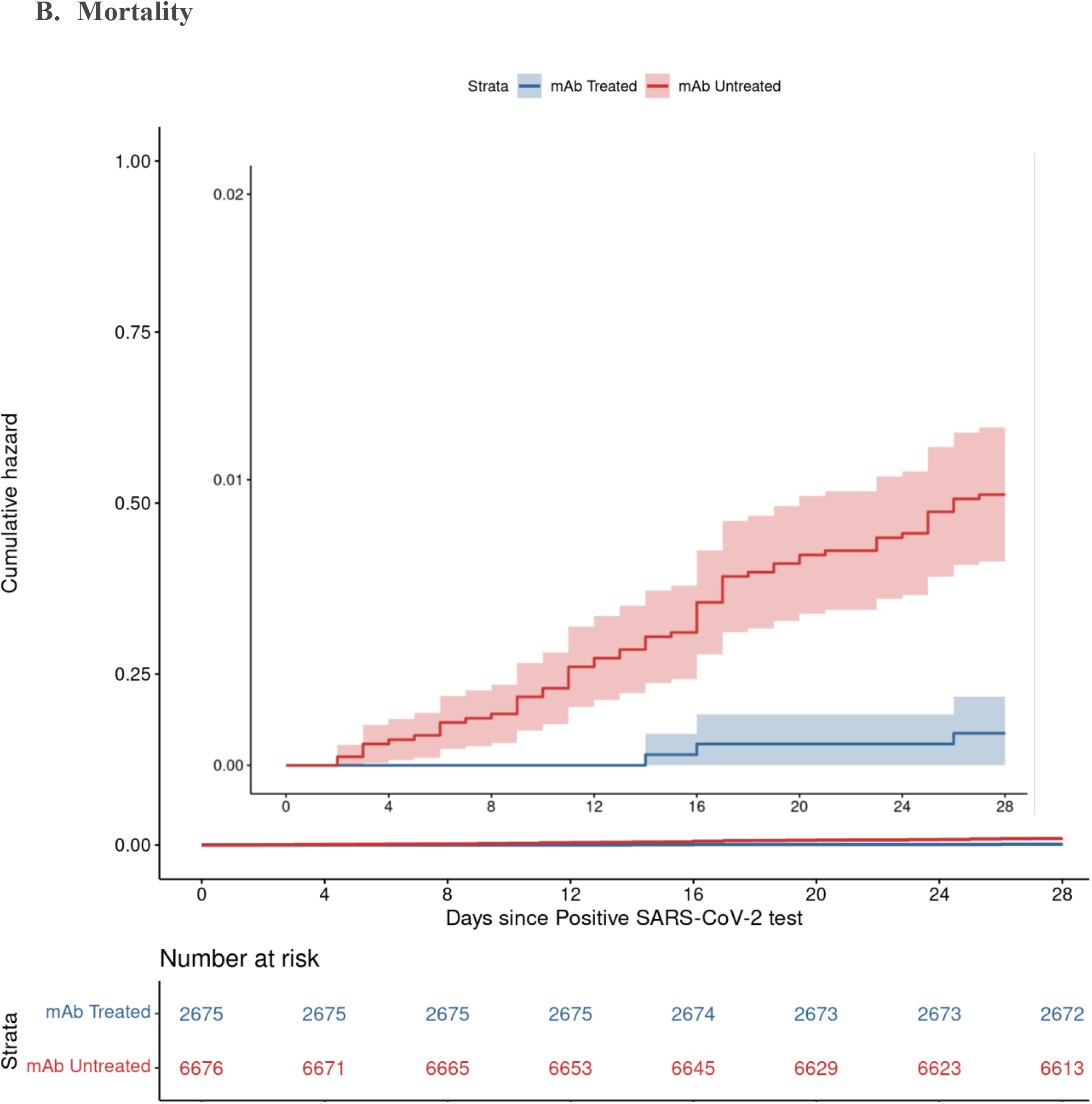
Cumulative Incidence Plots for All-Cause Hospitalization (A) and Mortality (B) to Day 28 by Monoclonal Antibody Treatment Status. A. Hospitalization B. Mortality

### Hospitalization and Mortality

The rate of 28-day all-cause hospitalization was lower among mAb-treated compared to matched mAb-untreated controls (4.0% v 7.7%; adjusted OR 0.48, 95%CI: 0.38-0.60) (**Table 2**; full model results **Appendix Table 3** in the Supplement). All-cause 28-day mortality in the mAb-treated group was 0.1% compared to 0.9% among the mAb-untreated group (adjusted OR 0.11, 95%CI: 0.03-0.29). These differences persisted to day 90 (adjusted OR 0.53; 95%CI: 0.44-0.65 for 90-day hospitalization and 0.17; 95%CI: 0.06-0.35 for 90-day mortality). Overall ED visit rates were higher for mAb-treated compared to mAb-untreated patients (18.7% vs. 16.9%; adjusted OR 1.24; 95%CI: 1.09-1.40); however, mAb-treated patients had fewer ED visits resulting in hospitalization (16.0% vs. 37.6%; adjusted OR 0.29, 95%CI: 0.21-0.38).

Based on a time-to-event analysis, the benefits associated with reduced hospitalization are largely accrued within 10 days of the positive test date, while the mortality benefit of mAb treatment continues to accrue over 28 days (**Figure 1**). Treatment benefits persisted to day 90 for both hospitalization and death (**Appendix Figure 2** in the Supplement).

**Figure 2.**
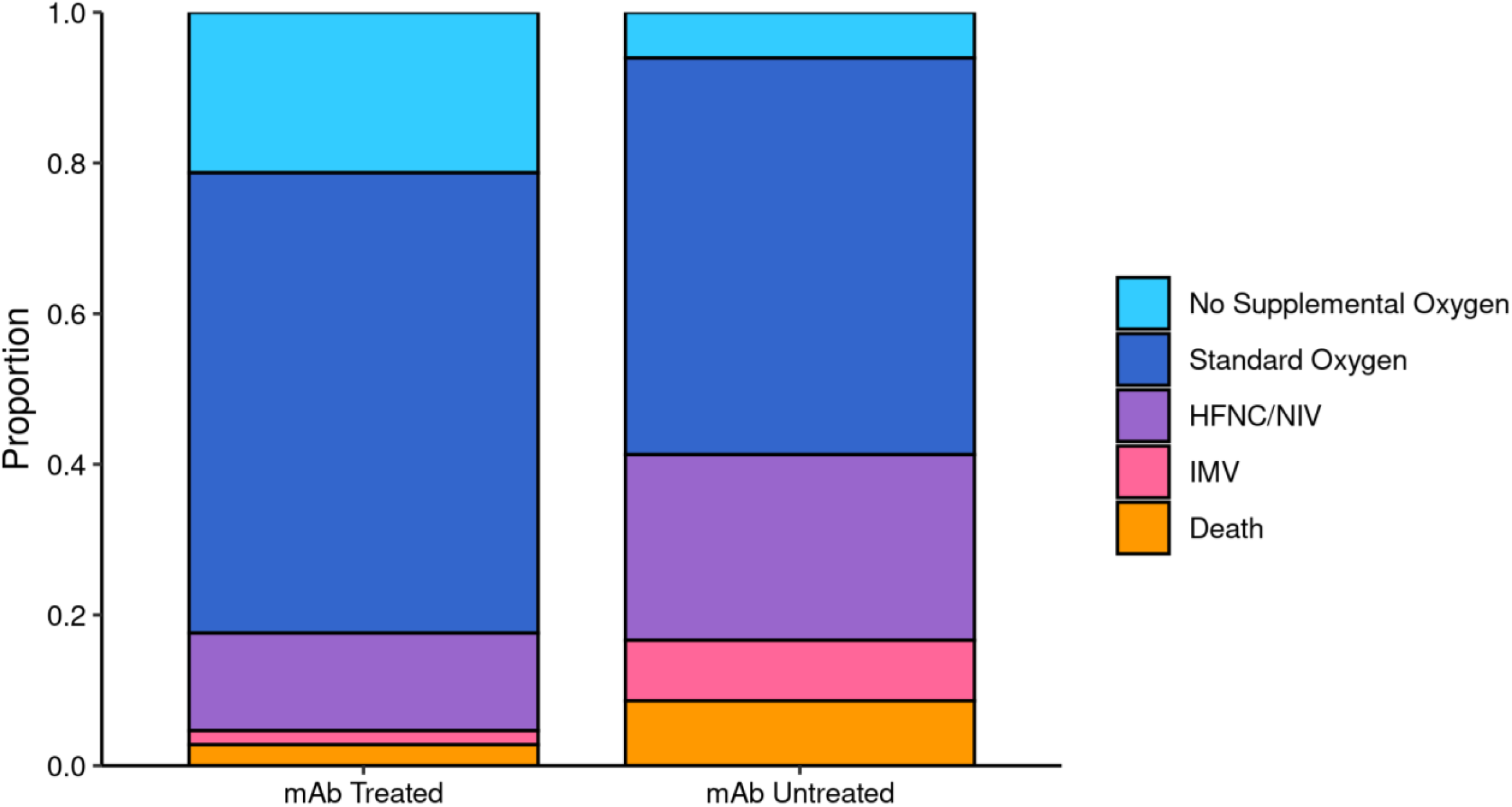
Maximum Respiratory Support by Monoclonal Antibody Treatment Status among Patients Hospitalized within 28 Days. Comparing severity of hospitalizations for n=108 mAb-treated and n=511 mAb-untreated patients, the maximum level of respiratory support was lower for mAb-treated patients (adjusted proportional OR 0.25; 95%CI: 0.16-0.38).

### Severity of Hospitalization

For patients requiring hospitalization, prior receipt of mAbs was associated with lower hospital LOS among survivors (5.8 vs. 8.5 days, adjusted incidence rate ratio 0.64, 95%CI: 0.51-0.82) and a lower rate of ICU admission (12.0% vs. 19.6%; adjusted OR 0.52, 95%CI 0.26-0.97), and mechanical ventilation or death (4.6% vs. 16.6%; adjusted OR 0.22, 95%CI: 0.07-0.52) (**Table 2**). For those requiring ICU care, prior receipt of mAbs was associated with shorter ICU LOS (3.5 vs. 8.6 days; adjusted incidence rate ratio 0.22; 95%CI: 0.10-0.48). Overall, severity of hospitalization was lower across the illness continuum for mAb-treated patients (**Figure 2**).

### Subgroup Analyses

The relative benefit of mAb therapy on reducing 28-day hospital admissions among key demographic and clinical subgroups was broadly similar across all subgroups (**Figure 3**). Of note, the association between mAb treatment and prevention of hospitalizations was at least as high during the Delta phase (OR 0.35; 95%CI: 0.25-0.50), compared to the Alpha phase (OR 0.67; 95%CI: 0.46-0.98). In addition, there was similar relative effectiveness for fully vaccinated (OR 0.44; 95%CI: 0.25-0.77) and not fully vaccinated (OR 0.49; 95%CI: 0.39-0.62) patients. However, the absolute treatment effect was higher for subgroups with higher baseline risk of hospitalization. For example, the number needed to treat (NNT) to prevent one hospitalization was 15 for patients age 65 years or older, 17 for those with at least 2 comorbid conditions, and 24 for those not fully vaccinated against SARS-CoV-2, compared to NNT of 45 for age 18-45 years, 88 for those without comorbidities, and 81 for fully vaccinated patients. Notably, only a small proportion of patients who were fully vaccinated against SARS-CoV-2 were hospitalized (1.8% of mAb-treated and 3.0% of mAb-untreated; **Figure 3**), and no patients died who were fully vaccinated and received mAb treatment.

**Figure 3.**
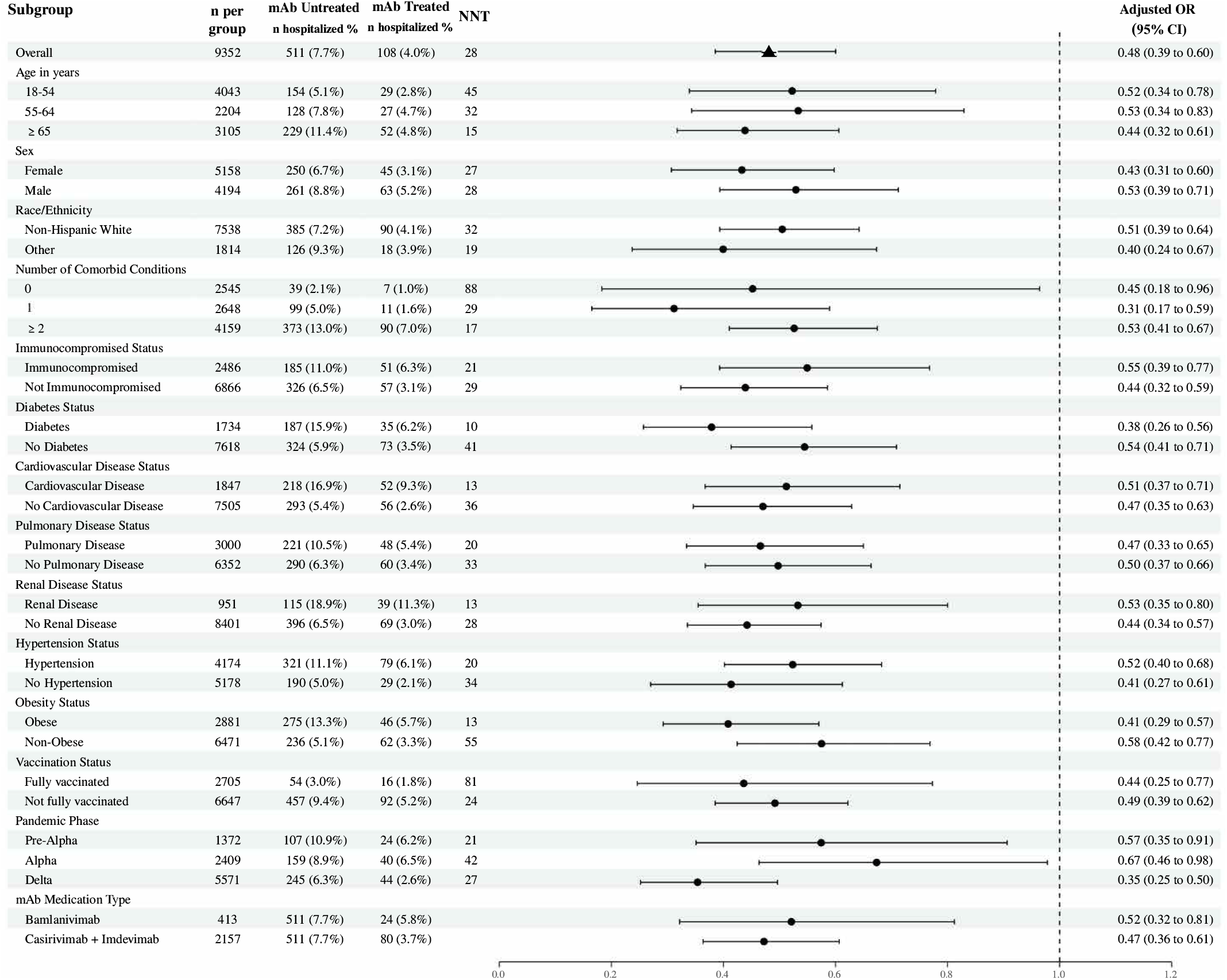
Subgroup Analysis of Monoclonal Antibody Effect on 28-day Hospitalization. For each subgroup, we calculated unadjusted rates of hospitalization, number needed to treat (NNT) to prevent one hospitalization (based on absolute risk reduction in unadjusted hospitalization rates), and adjusted relative odds of hospitalization. Each adjusted odd ratio represents a separate model. All regression models adjusted for age, sex, race/ethnicity, BMI, immunocompromised status, number of comorbidities, insurance status, pandemic phase, and vaccination status. Results were not adjusted for multiple comparisons. Abbreviations: mAb, monoclonal antibody; NNT, number needed to treat; OR, odds ratio; CI, confidence interval

### Sensitivity Analyses

Two sensitivity analyses were performed, the first restricting the cohort to only patients meeting EUA eligibility criteria based on available EHR data and the second using a more conservative imputation method when the date of positive SARS-CoV-2 test was missing. Neither analyses materially changed the main results (**Appendix Tables 4-7** in the Supplement).

## DISCUSSION

We report real-world evidence that demonstrates novel results on both high effectiveness of mAb treatment in reducing hospitalization during the Delta variant phase and a remarkable overall mortality benefit with an 89% lower mortality at 28 days. Neutralizing mAbs are widely seen as important tools for managing surging cases of COVID-19, yet prior studies could not evaluate effectiveness of mAbs against Delta variant infections and have been underpowered to evaluate impact of mAbs on the most clinically important outcome: patient mortality. The present study fills these key knowledge gaps.

There have also been critical gaps in understanding the effects of mAbs on important subgroups of patients, such as those with older age, comorbid conditions, and prior SARS-CoV-2 vaccination. With our large sample size, we demonstrated clinical benefits of mAb administration among virtually all subgroups examined, with similar relative benefits in terms of reduced odds of hospitalizations across all subgroups. These subgroup findings highlight the need to interpret relative benefits in light of highly variable absolute hospitalization rates, because the NNT to avert one hospitalization depends on both mAb effectiveness and baseline rates of hospitalization. For example, we found a similar relative effect size for vaccinated and unvaccinated patients, but the NNT to avert one hospitalization among unvaccinated patients is 24, while the NNT for vaccinated patients is 81. These results are of practical importance for policymakers and clinicians because there have been shortages of mAb supplies and infusion capacity.^11,12^ Specifically, our findings suggest the most efficient use of limited mAb infusion capacity to alleviate strain on hospitals is to preferentially administer mAbs to patients at highest baseline risk for hospitalization, including those who are older, not fully vaccinated, or with multiple comorbid conditions. Notably, 28-day hospitalization among mAb-treated but not fully vaccinated patients was almost 3-fold higher (5.2%) than for mAb-treated patients who were fully vaccinated (1.8%) and higher even than mAb-untreated patients who were fully vaccinated (3.0%). These data support that SARS-CoV-2 vaccination remains the first line intervention to prevent COVID-19 hospitalizations with mAb treatment best used as supplemental therapy for high-risk patients.

### Limitations

This study has several limitations. The setting was a single health system; while large and representing both urban and rural settings and community and academic hospitals, it is geographically limited to one US state. Our sample had relatively low racial and ethnic minority representation, limiting our ability to detect differences across these key subgroups. While we used statewide data for mortality and vaccination status, hospitalizations were collected only within this single health system. If mAb-untreated patients were also less likely to be seen in the health system for other services (hence, more likely to be hospitalized elsewhere), this may bias our results toward the null. We also relied on EHR data, including manual chart reviews, which may have missing or inaccurate information about the presence of chronic conditions.^21^ These factors might have limited our ability to detect the impact of mAb treatment, especially between subgroups. Our EHR data does not contain information on SARS-CoV-2 variants at the patient level, so variant phases are presented chronologically. However, during Colorado’s Delta phase more than 99% of sequenced SARS-CoV-2 was Delta variant.^22^ Our large sample size allowed the detection of meaningful benefits of mAb therapy for most subgroups, but the study could not detect potentially relevant differences between subgroups. Our propensity scoring method achieved excellent matching between mAb-treated and mAb-untreated patient groups across multiple variables, but unmeasured confounders may remain. Finally, our study was conducted prior to the emergence of the Omicron variant and there is in vitro evidence of reduced SARS-CoV-2 neutralization by some authorized mAbs.^23,24^ Forthcoming studies will evaluate the effectiveness of each available mAb treatment during the Omicron phase of the pandemic.

### Conclusion

Using real-world evidence, this study demonstrated effectiveness of mAb treatment in reducing hospitalizations among COVID-19 outpatients, including during the Delta variant phase, as well as a remarkable 89% overall reduction in mortality at 28 days, compared to matched mAb-untreated patients. For hospitalized patients, prior mAb treatment was associated with notably lower disease severity, including reduced hospital length of stay, ICU length of stay, mechanical ventilation, and death. When access to mAbs is limited, prioritizing patients at highest risk for hospitalization has the most potential to reduce health system strain during the COVID-19 pandemic.

## Data Availability

All data produced in the present study may be made available upon reasonable request to the corresponding author.

## ACKNOWLEDGMENTS

This study was funded by National Institutes of Health / National Center for Advancing Translational Sciences grants UL1TR002525, UL1TR002535-03S3 and UL1TR002535-04S2. Dr. Wynia received research funding from PCORI and ASPR and is an unpaid advisor to NASEM, including on crisis standards of care during the COVID-19 pandemic, and to DARPA, the Hastings Center, and the Lancet on projects unrelated to mAbs. Dr. Bennett received research grants from the NIH outside of the current work. Dr. Carlson received research grants from the NIH outside the current work. Dr. Ginde received other COVID-19 research grants from NIH, DoD, CDC, AbbVie, and Faron Pharmaceuticals, outside the current work. Other authors have no disclosures to report.

