## Supplementary Appendix for "Real World Evidence of Neutralizing Monoclonal Antibodies for Preventing Hospitalization and Mortality in COVID-19 Outpatients"

This Appendix has been provided by the authors to give readers additional information about the work.

### Monoclonal Antibody (mAb) Colorado Research Team and Collaborators

University of Colorado Anschutz Medical Campus, Aurora, CO

***Project Leadership and Administration***: Adit A. Ginde, MD, MPH; Ronald J. Sokol, MD; Tim Lockie, MS, MBA; Heather R. Stocker, MA; Mujeeb Zaird, MBA

***Biostatistics, Epidemiology, and Research Design Core:*** Nichole E. Carlson, PhD; Laurel E. Beaty, MS; Adane F. Wogu, PhD; David A. Mayer, BS; Samantha Roberts, MPH; Megan Branda, MS

***Clinical Core***: Matthew K. Wynia, MD, MPH; Julie G. Ressalam, MPH; Matthew DeCamp, MD, PHD**;** Hilary Lum, MD, PhD; Matthew A. Miller, PharmD, BCP, BCIDP; Kyle Molina, PharmD, BCIDP; Ruthvik Reddy Perikiti, MS; Lauren Shviraga Whitesitt, BS

***Informatics Core***: Tellen D. Bennett, MD, MS; Toan C. Ong, PhD; Seth Russell, MS; Jeffrey Steele, RN; Margaret Rebull, MA; Ian Brooks, PhD; Madelyne Hull, MPH; Aaron Mobley, PhD; Supported by the Health Data Compass Data Warehouse project team and Clinical Research Support Team (CReST)

***Dissemination and Implementation Core***: Bethany M. Kwan, PhD, MSPH; Vanessa Owen, MA; Chelsea Sobczak, MPH; Jenna Reno, PhD; Mika Hamer, MPH, PhD; Eric G. Campbell, PhD

University of Colorado Health (UCHealth) System

Richard D. Zane, MD; Christopher B. Davis, MD; Kathy Deanda, RN, MSN; Mathew Miller, PharmD; Kyle Molina, PharmD

Colorado Department of Public Health and Environment, Denver, CO

Eric France, MD, MSPH, MBA; Wendy M. Bamberg, MD; Alexis W. Burakoff, MD, MPH; Diana M. Tapay, MA; Shannon O'brien, MD, MPH; Amanda Hettinger, MA; Rachel Severson, MS; Elaine M. Sabyan

Tri-County Health Department

John M. Douglas, Jr, MD; Alix Rizzuto, MS, RN; Samantha Smith, MS, RDN

We also wish to thank patients and families for their participation in research to accelerate discovery and rapidly advance clinical care in the pandemic; numerous colleagues who provided support for this project; frontline health care workers for their tireless efforts and life-saving contributions; and researchers around the world working together in the quest to inform healthcare practice and improve patient outcomes of COVID-19.

### Supplementary Methods

#### *Description of Demographic Variables*

Age was determined at the time of positive SARS-CoV-2 test or mAb administration if SARS-CoV-2 test date was not available in the electronic health record (EHR). We categorized age into 18-54, 55-64, and ≥65 years, based on thresholds that were defined in the original monoclonal antibody (mAb) Emergency Use Authorization (EUA) criteria (Table S8). Sex was defined as legal sex in the EHR and was binarized into female and male (non-binary status was not explicitly defined), and this field was missing for two subjects. To preserve sample size, the variables race and ethnicity were combined and categorized into non-Hispanic white, non-Hispanic black, Hispanic, and other. In the subgroup analysis race/ethnicity is binarized into non-Hispanic white and other to allow for a large enough sample size for subgroup analyses. Continuous body mass index (BMI in $kg/m^2$) was categorized into 4 categories: underweight (<18.5), normal weight (18.5-24.9), overweight (25.0-29.9), obese (≥30.0). The number of comorbid medical conditions was calculated using obesity, hypertension, cardiovascular disease, diabetes, pulmonary disease, and renal disease, and was categorized into none, one, or two or more. Immunocompromised status was categorized separately. All individual comorbid conditions were considered as binary variables with either evidence of the comorbid condition or no evidence of the comorbid condition in the EHR.

#### *EHR Curation of Comorbidities*

We defined comorbidities based on the updated Charlson and Elixhauser Comorbidity Indices[1,2] as implemented in the 'icd' R package[3] and reported previously from the same health system.[4] From the eligibility criteria above, we categorized a patient as having diabetes, renal disease, pulmonary disease, cardiovascular disease, or immunocompromised status if those conditions were present for that patient in either the Charlson or Elixhauser system. For obesity and hypertension, we used only the Elixhauser system. Because of the importance of immunocompromised status as a risk factor for hospitalization and mortality from COVID-19, we additionally defined patients as immunocompromised if any of the below medications were present in the EHR medication administration record during the 90-day lookback period. The list of medications was developed jointly by an expert team of UCHealth pharmacists and Infectious Disease physicians. We evaluated the accuracy of the EHR medication curation by manually reviewing the charts of 2,555 patients. EHR curation accurately classified 85% of patients. Potentially discordant patients most often had received immune-suppressing medications prior to our IRB-approved 90-day lookback period or had received prednisone or methylprednisolone at doses under the expert-defined dose threshold.

*List of immune-suppressing medications*

- Alemtuzumab
- Belatacept in past 2 months
- Calcineurin inhibitors (tacrolimus and cyclosporine – excludes topical/ophthalmic administration routes)
- Eculizumab
- mTOR-inhibitors (everolimus, sirolimus – excludes topical routes)
- Mycophenolate, azathioprine, cyclophosphamide in the last 1 month
- Prednisone or methylprednisolone, oral or IV only (≥10 mg prednisone equivalent)
- Rituximab

- Thymoglobulin
- TNF-α inhibitor (e.g., infliximab, etanercept, golimumab, adalimumab, certolizumab)

***Missing Data Techniques***

Of the 3,164 patients who received mAb treatment, 1,593 (50.3%) were missing an initial positive SARS-CoV-2 test date in the UCHealth EHR, suggesting many initial tests were performed outside the UCHealth system. For the primary analysis, a distribution of the time difference between positive SARS-CoV-2 test date and mAb administration date was created for subjects who had both. Then, time differences were randomly sampled with replacement from this distribution and were used to impute positive test dates for the patients who only had a mAb administration date. We evaluated a sensitivity analysis to this approach by imputing the maximum allowed time difference between SARS-CoV-2 positive date and mAb administration date (10 days) for all patients missing the first date.

In the full cohort (prior to propensity matching), 20,010 (55.5%) of patients were missing BMI. This is typical of EHR studies. A missing category for BMI was introduced and BMI with 5 levels (<18.5, 18.5-24.9, 25.0-29.9, ≥30.0 kg/m$^2$, and missing) was used during propensity matching and analysis. A combination of reported BMI and reported obesity status was used in determining eligibility. A patient was considered eligible if they had a reported BMI higher than the threshold (either 25 or 35 depending on the date) or if they were indicated as "obese" in the EHR.

A complete case analysis was performed for propensity matching. All comorbid conditions were missing from the EHR for 2,077 (5.7%) patients. Race/ethnicity was missing for 1,996 (5.5%) of patients. A total of 3,407 (9.4%) of patients were removed for the propensity matching. Although there are limitations to a complete case analysis, the ability to accurately impute missingness based on the available EHR variables was limited. Thus, the potential bias of a complete case analysis for this amount of missingness was believed to be smaller than the potential bias from a small imputation model.

***Propensity Matching***

The propensity matched dataset was created through a logistic regression propensity score matching process. Nearest neighbor matching was used, with a maximum ratio of 3:1 mAb-untreated and mAb-treated groups. In the matching process, we lost both mAb-treated and mAb-untreated subjects and ended up with a ratio of approximately 2.5:1. A common support was used for both the cases and controls, and a caliper width of < 0.2*SD of the propensity distribution was applied[4]. The standardized mean differences of each level of all covariates included in the model were calculated to compare the means and prevalence in the propensity matched dataset. A standardized mean difference of <0.1 was considered to have a non-meaningful imbalance in the data[3].

The baseline characteristics included in the propensity matching process were age in years, sex, race/ethnicity, BMI, insurance status, immunocompromised status, number of other comorbid conditions, and days from initial cohort date, November 20, 2020 (as a quadratic effect).

***Model Fitting***

Each of the models presented in Table 2 were fitted using the same group of adjustment variables. The variables that were included were age, sex, race/ethnicity, BMI, insurance status, vaccination status, pandemic phase, number of comorbid conditions, and immunocompromised status. A significance level of 0.05 was used to determine statistical significance; 95% confidence intervals (CIs) were also used to evaluate clinical significance.

#### *Subgroup Analysis*

To evaluate the potential heterogeneity of the treatment effect across key subgroups of interest the above model for the primary outcome (28-day hospitalization) was fitted separately for each of the 14 subgroups of interest. More specifically, for each subgroup an interaction was included between the subgroup variable and the treatment variable, and the main effects of the other variables included for adjustment. The subgroups investigated included age in years, sex, race/ethnicity, number of comorbid conditions, immunocompromised status, diabetes status, cardiovascular disease status, pulmonary disease status, renal disease status, hypertension status, obesity status, vaccination status, pandemic phase, and mAb medication type.

A total of 29 treatment effects were estimated and all subgroup analyses that were performed were reported in Figure 3. Heterogeneity was assessed visually through a forest plot, and subgroup odds ratio estimates were compared. Statistical tests of significance for each interaction term were not reported in the paper, as this analysis was likely underpowered and the potential for Type I error due to multiple comparisons was not accounted for[4]. Subgroup analyses were post hoc specified. Within-level results are presented for each estimated treatment effect by subgroup.

In addition, raw counts and rates are reported for each subgroup. The number needed to treat (NNT) was calculated based on the raw rates as the inverse of the absolute risk reduction. NNT was not calculated for mAb medication type.

### Appendix Figures

#### Appendix Figure 1: Flow of Patients into the Primary Study Cohort

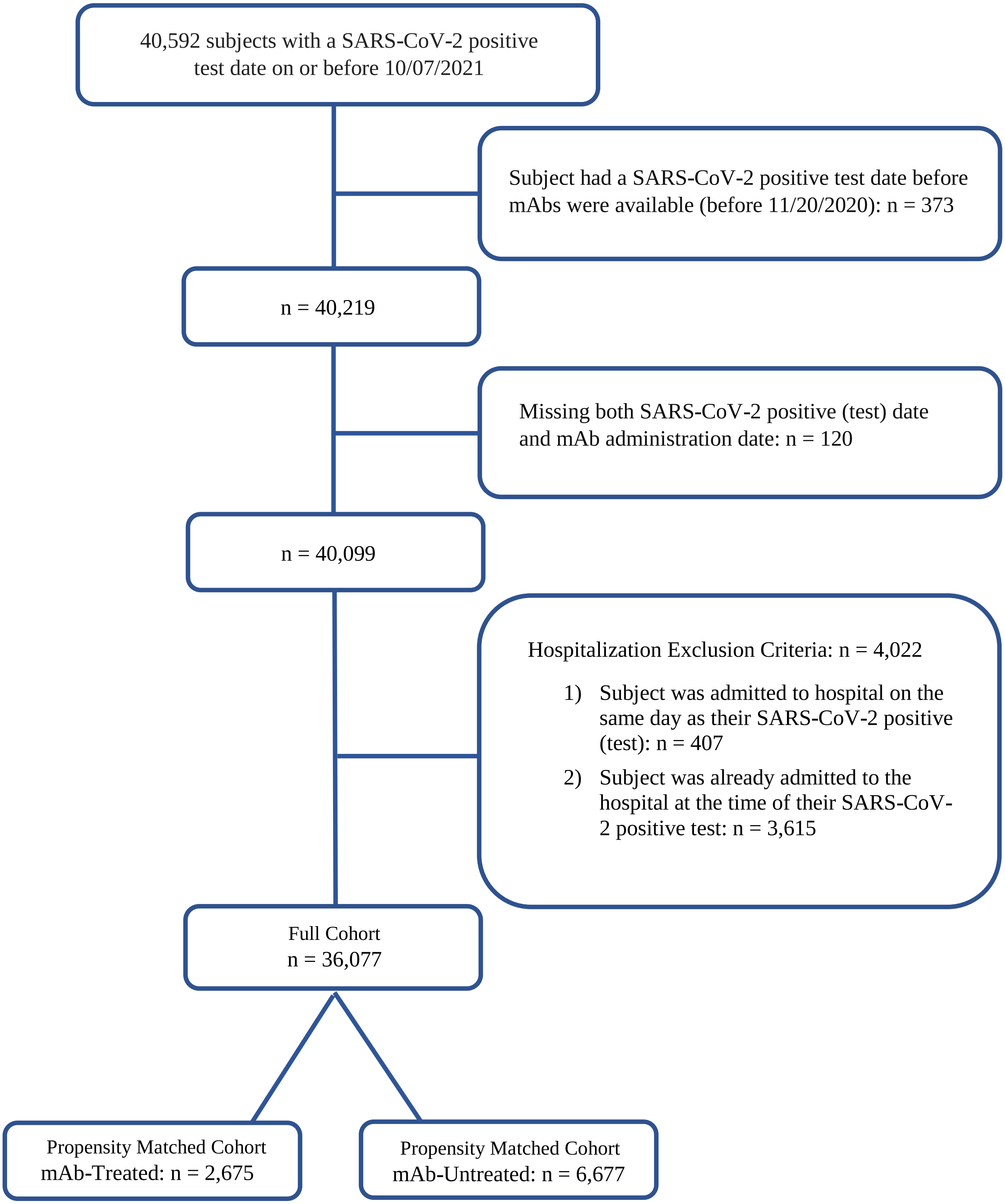

**Appendix Figure 2. Cumulative Incidence Plots for All-Cause Hospitalization (A) and Mortality (B) to Day 90 by Monoclonal Antibody Treatment Status**

A.

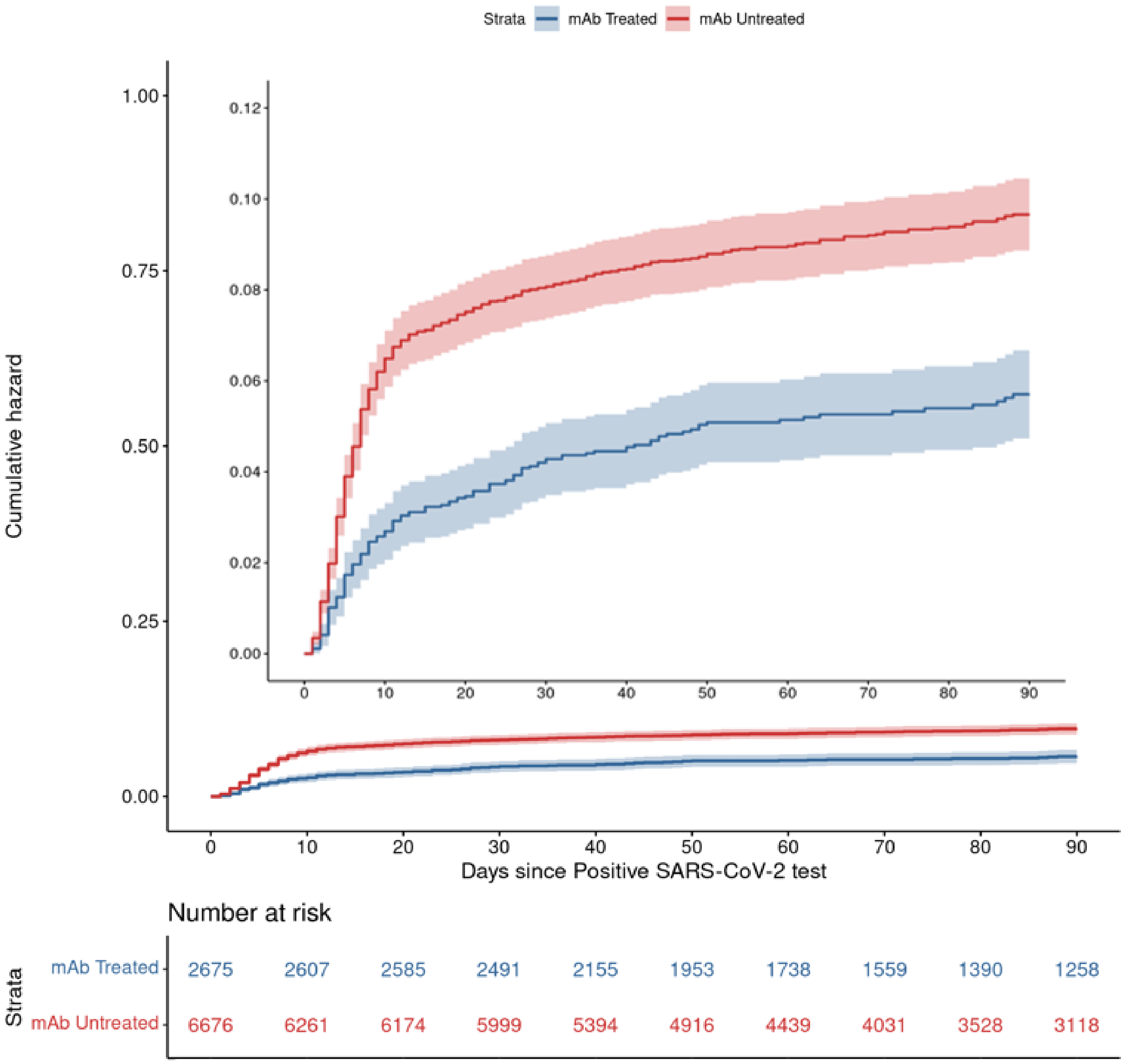

B.

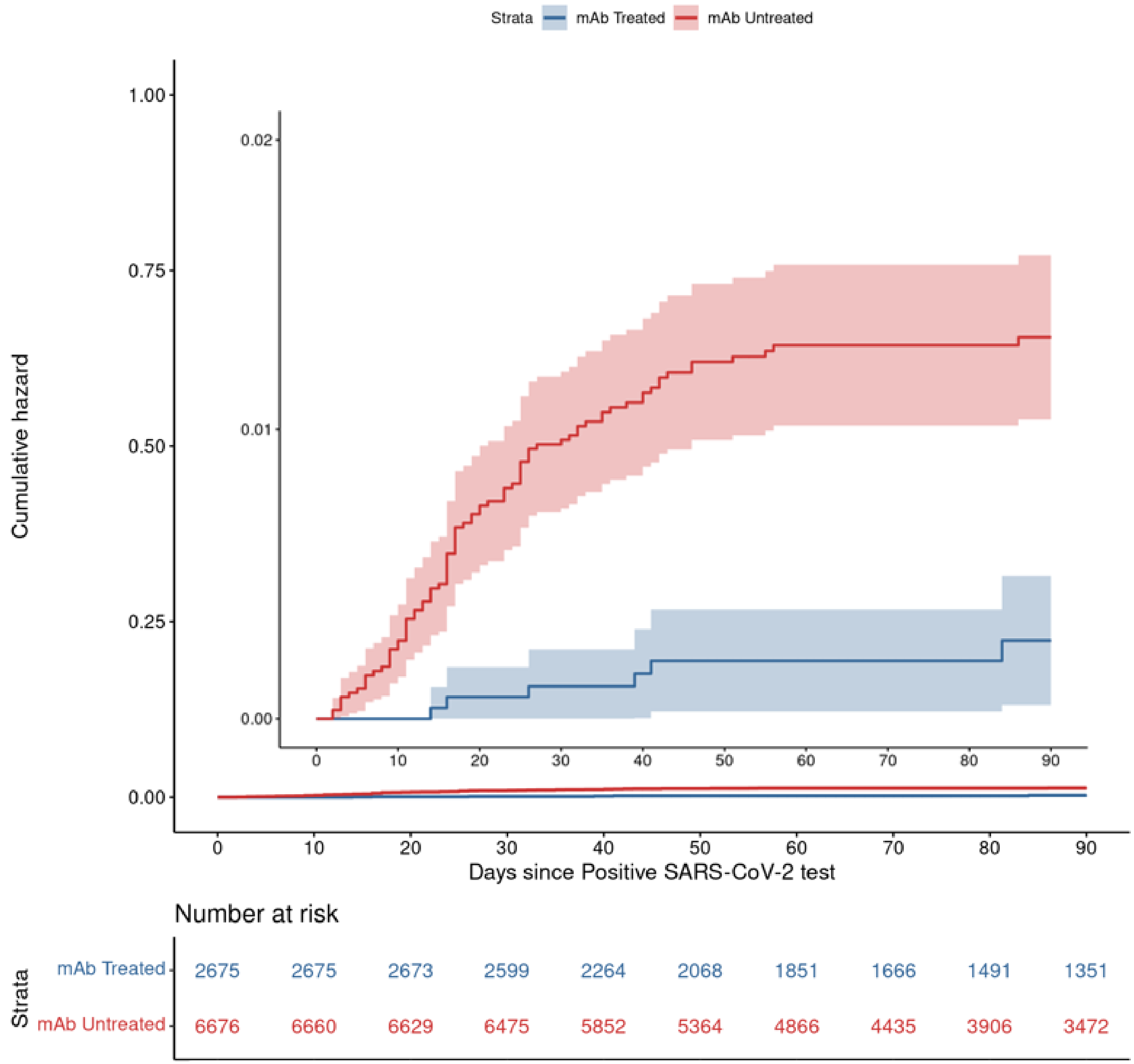

Strata
mAb Treated
mAb Untreated
Cumulative hazard
1.00
0.75
0.50
0.25
0.00
0.02
0.01
0.00
0
10
20
30
40
50
60
70
80
90
Days since Positive SARS-CoV-2 test
Number at risk
Strata
mAb Treated
mAb Untreated
2675 2675 2673 2599 2264 2068 1851 1666 1491 1351
6676 6660 6629 6475 5852 5364 4866 4435 3906 3472

### Appendix Tables

**Appendix Table 1. Baseline Characteristics by Monoclonal Antibody Treatment Status for Full SARS-CoV-2 Positive Cohort, Prior to Propensity Matching**

| Characteristic | mAb-Treated n=2758 | mAb-Untreated n=33319 |
|---|---|---|
| Age in years | | |
| 18-54 years | 1052 (38.1%) | 25075 (75.3%) |
| 55-64 years | 586 (21.2%) | 4669 (14.0%) |
| ≥65 years | 1120 (40.6%) | 3575 (10.7%) |
| Sex | | |
| Female | 1491 (54.1%) | 17681 (53.1%) |
| Male | 1267 (45.9%) | 15636 (46.9%) |
| Missing | 0 (0.0%) | 2 (0.0%) |
| Race/Ethnicity | | |
| Non-Hispanic White | 2229 (80.8%) | 22311 (67.0%) |
| Hispanic | 267 (9.7%) | 5263 (15.8%) |
| Non-Hispanic Black | 64 (2.3%) | 1488 (4.5%) |
| Other | 133 (4.8%) | 2326 (7.0%) |
| Missing | 65 (2.4%) | 1931 (5.8%) |
| Body mass index in kg/m$^2$ | | |
| <18.5 | 23 (0.8%) | 204 (0.6%) |
| 18.5-24.9 | 365 (13.2%) | 3784 (11.4%) |
| 25.0-29.9 | 577 (20.9%) | 4666 (14.0%) |
| ≥30.0 | 780 (28.3%) | 5668 (17.0%) |
| Missing | 1013 (36.7%) | 18997 (57.0%) |
| Immunocompromised | | |
| Yes | 819 (29.7%) | 3281 (9.8%) |
| No | 1917 (69.5%) | 27993 (84.0%) |
| Missing | 22 (0.8%) | 2045 (6.1%) |
| Number of Other Comorbid Conditions | | |
| 0 | 747 (27.1%) | 18488 (55.5%) |
| 1 | 689 (25.0%) | 6887 (20.7%) |
| ≥2 | 1299 (47.1%) | 5890 (17.7%) |
| Missing | 23 (0.8%) | 2054 (6.2%) |
| Diabetes | | |
| Yes | 567 (20.6%) | 2523 (7.6%) |
| No | 2168 (78.6%) | 28742 (86.3%) |
| Missing | 23 (0.8%) | 2054 (6.2%) |
| Cardiovascular Disease | | |
| Yes | 563 (20.4%) | 2346 (7.0%) |
| No | 2172 (78.8%) | 28919 (86.8%) |
| Missing | 23 (0.8%) | 2054 (6.2%) |
| Pulmonary Disease | | |
| Yes | 896 (32.5%) | 5628 (16.9%) |

| | | |
|---|---|---|
| No | 1839 (66.7%) | 25637 (76.9%) |
| Missing | 23 (0.8%) | 2054 (6.2%) |
| Renal Disease | | |
| Yes | 349 (12.7%) | 1108 (3.3%) |
| No | 2386 (86.5%) | 30157 (90.5%) |
| Missing | 23 (0.8%) | 2054 (6.2%) |
| Hypertension | | |
| Yes | 1310 (47.5%) | 6321 (19.0%) |
| No | 1425 (51.7%) | 24944 (74.9%) |
| Missing | 23 (0.8%) | 2054 (6.2%) |
| Obesity | | |
| Yes | 814 (29.5%) | 5089 (15.3%) |
| No | 1921 (69.7%) | 26176 (78.6%) |
| Missing | 23 (0.8%) | 2054 (6.2%) |
| Vaccination Status | | |
| Not known to be vaccinated | 1678 (60.8%) | 28250 (84.8%) |
| Partially vaccinated | 154 (5.6%) | 1630 (4.9%) |
| Fully vaccinated | 926 (33.6%) | 3439 (10.3%) |
| Pandemic Phase | | |
| Pre-Alpha: Nov 2020 - Feb 2021 | 399 (14.5%) | 16926 (50.8%) |
| Alpha: March 2021 - June 2021 | 628 (22.8%) | 7995 (24.0%) |
| Delta: July 2021 - Sep 2021 | 1731 (62.8%) | 8398 (25.2%) |
| Type of Monoclonal Antibody | | |
| Bamlanivimab | 425 (15.4%) | - |
| Bamlanivimab + etesevimab | 95 (3.4%) | - |
| Casirivimab + imdevimab | 2220 (80.5%) | - |
| Sotrovimab | 18 (0.7%) | - |

Abbreviations: mAb, monoclonal antibody; SD, standard deviation

**Appendix Table 2. Standard Mean Differences for Propensity Matched Cohort**

| Characteristic | mAb-Treated Mean | mAb-Untreated Mean | Standardized Mean Difference |
|---|---|---|---|
| Distance | 0.234 | 0.231 | 0.014 |
| Age in years | | | |
| 18-54 years | 0.381 | 0.385 | -0.009 |
| 55-64 years | 0.213 | 0.246 | -0.081 |
| ≥65 years | 0.407 | 0.369 | 0.076 |
| Sex | | | |
| Female | 0.543 | 0.552 | -0.018 |
| Male | 0.457 | 0.448 | 0.018 |
| Race/Ethnicity | | | |
| Non-Hispanic White | 0.099 | 0.107 | -0.029 |
| Hispanic | 0.024 | 0.025 | -0.008 |
| Non-Hispanic Black | 0.828 | 0.813 | 0.039 |
| Other | 0.049 | 0.054 | -0.023 |
| Body mass index, $kg/m^2$ * | | | |
| <18.5 | 0.009 | 0.009 | -0.004 |
| 18.5-24.9 | 0.135 | 0.136 | -0.003 |
| 25.0-29.9 | 0.213 | 0.216 | -0.007 |
| ≥30.0 | 0.288 | 0.302 | -0.032 |
| Missing | 0.355 | 0.336 | 0.040 |
| Immunocompromised | | | |
| Immunocompromised | 0.698 | 0.710 | -0.028 |
| Not immunocompromised | 0.302 | 0.290 | 0.028 |
| Number of Other Comorbid Conditions | | | |
| 0 | 0.265 | 0.243 | 0.050 |
| 1 | 0.255 | 0.278 | -0.053 |
| ≥2 | 0.481 | 0.480 | 0.002 |
| Insurance | | | |
| Private/Commercial | 0.061 | 0.071 | -0.039 |
| Medicare | 0.393 | 0.361 | 0.066 |
| Medicaid | 0.016 | 0.015 | 0.010 |
| None/Uninsured | 0.022 | 0.025 | -0.016 |
| Other/Unknown | 0.507 | 0.528 | -0.044 |
| Time between Cohort Inception (11/20/2020) and SARS-CoV-2 test | | | |
| Days | 223.219 | 223.48 | -0.003 |
| $Days^2$ | 57946.6 | 57682.9 | 0.008 |

Abbreviations: mAb, monoclonal antibody; SAR-CoV-2, severe acute respiratory syndrome coronavirus-2

### Appendix Table 3. Full Model Results for 28-Day Hospitalization Primary Outcome

| Characteristic | Adjusted OR | 95% CI |
|---|---|---|
| Treatment Status | | |
| mAb-Untreated | Reference | |
| mAb-Treated | 0.48 | (0.38, 0.60) |
| Age in years | | |
| 18-54 | Reference | |
| 55-65 | 1.42 | (1.12, 1.80) |
| ≥65 | 1.37 | (1.00, 1.89) |
| Sex | | |
| Female | Reference | |
| Male | 1.51 | (1.27, 1.80) |
| Race/Ethnicity | | |
| Non-Hispanic White | Reference | |
| Hispanic | 1.30 | (1.00, 1.67) |
| Non-Hispanic Black | 0.72 | (0.40, 1.21) |
| Other | 1.57 | (1.08, 2.25) |
| Insurance Status | | |
| Private/Commercial | Reference | |
| Medicare | 1.57 | (1.17, 2.10) |
| Medicaid | 1.22 | (0.87, 1.69) |
| None/Uninsured | 1.28 | (0.51, 2.72) |
| Other/Unknown | 1.17 | (0.63, 2.03) |
| Body Mass Index in $kg/m^2$ | | |
| <18.5 | Reference | |
| 18.5-24.9 | 1.13 | (0.50, 2.28) |
| 25.0-29.9 | 0.89 | (0.69, 1.15) |
| ≥30.0 | 0.91 | (0.71, 1.17) |
| Missing | 0.03 | (0.01, 0.06) |
| Immunocompromised Status | | |
| No | Reference | |
| Yes | 1.22 | (1.01, 1.46) |
| Number of Other Comorbid Conditions | | |
| 0 | Reference | |
| 1 | 1.66 | (1.17, 2.40) |
| ≥2 | 3.78 | (2.73, 5.34) |
| Pandemic Phase | | |
| Pre-Alpha | Reference | |
| Alpha | 1.39 | (1.08, 1.79) |
| Delta | 1.51 | (1.18, 1.95) |
| Vaccination Status | | |
| Fully vaccinated | Reference | |
| Not known to be vaccinated | 3.65 | (2.77, 4.87) |

Abbreviations: mAb, monoclonal antibody; OR, odds ratio; CI, confidential interval

**Appendix Table 4.** **Baseline Characteristics by Monoclonal Antibody Treatment Status for Sensitivity Analysis Cohort 1**

| Characteristic | Full Cohort | | Matched Cohort | |
|---|---|---|---|---|
| | mAb-Treated (n=2497) | mAb-Untreated (n=14549) | mAb-Treated (n = 2445) | mAb-Untreated (n = 5943) |
| Age in years* | | | | |
| 18-54 | 870 (34.8%) | 8172 (56.2%) | 857 (35.1%) | 2566 (43.2%) |
| 55-64 | 507 (20.3%) | 2802 (19.3%) | 501 (20.5%) | 1346 (22.6%) |
| ≥65 | 1120 (44.9%) | 3575 (24.6%) | 1087 (44.5%) | 2031 (34.2%) |
| Sex* | | | | |
| Female | 1367 (54.7%) | 8195 (56.3%) | 1343 (54.9%) | 3266 (55.0%) |
| Male | 1130 (45.3%) | 6354 (43.7%) | 1102 (45.1%) | 2677 (45.0%) |
| Race/Ethnicity* | | | | |
| Non-Hispanic White | 2004 (80.3%) | 9447 (64.9%) | 1996 (81.6%) | 4694 (79.0%) |
| Hispanic | 258 (10.3%) | 2867 (19.7%) | 256 (10.5%) | 742 (12.5%) |
| Non-Hispanic Black | 63 (2.5%) | 914 (6.3%) | 63 (2.6%) | 172 (2.9%) |
| Other | 131 (5.2%) | 1053 (7.2%) | 130 (5.3%) | 335 (5.6%) |
| Missing | 41 (1.6%) | 268 (1.8%) | - | - |
| Insurance Status* | | | | |
| Medicaid | 144 (5.8%) | 1830 (12.6%) | 1176 (48.1%) | 3302 (55.6%) |
| Medicare | 1073 (43.0%) | 3408 (23.4%) | 1047 (42.8%) | 1982 (33.4%) |
| None/Uninsured | 42 (1.7%) | 484 (3.3%) | 142 (5.8%) | 430 (7.2%) |
| Other/Unknown | 45 (1.8%) | 532 (3.7%) | 35 (1.4%) | 93 (1.6%) |
| Private/Commercial | 1193 (47.8%) | 8295 (57.0%) | 45 (1.8%) | 136 (2.3%) |
| Body Mass Index, kg/m$^2$* | | | | |
| <18.5 | 21 (0.8%) | 93 (0.6%) | 20 (0.8%) | 48 (0.8%) |
| 18.5-24.9 | 321 (12.9%) | 1590 (10.9%) | 318 (13.0%) | 729 (12.3%) |
| 25.0-29.9 | 569 (22.8%) | 2855 (19.6%) | 563 (23.0%) | 1292 (21.7%) |
| ≥30.0 | 772 (30.9%) | 4799 (33.0%) | 762 (31.2%) | 1893 (31.9%) |
| Missing | 814 (32.6%) | 5212 (35.8%) | 782 (32.0%) | 1981 (33.3%) |
| Immunocompromised* | | | | |
| Yes | 819 (32.8%) | 3281 (22.6%) | 808 (33.0%) | 1676 (28.2%) |
| No | 1666 (66.7%) | 10996 (75.6%) | 1637 (67.0%) | 4267 (71.8%) |
| Missing | 12 (0.5%) | 272 (1.9%) | - | - |
| Number of Other Comorbid Conditions* | | | | |
| 0 | 509 (20.4%) | 3557 (24.4%) | 492 (20.1%) | 1358 (22.9%) |
| 1 | 678 (27.2%) | 4993 (34.3%) | 670 (27.4%) | 1801 (30.3%) |
| ≥2 | 1297 (51.9%) | 5718 (39.3%) | 1283 (52.5%) | 2784 (46.8%) |
| Missing | 13 (0.5%) | 281 (1.9%) | - | - |
| Diabetes | | | | |
| Yes | 567 (22.7%) | 2523 (17.3%) | 561 (22.9%) | 1126 (18.9%) |
| No | 1917 (76.8%) | 11745 (80.7%) | 1884 (77.1%) | 4817 (81.1%) |
| Missing | 13 (0.5%) | 281 (1.9%) | - | - |
| Cardiovascular Disease | | | | |

| | | | | |
|---|---|---|---|---|
| Yes | 562 (22.5%) | 2264 (15.6%) | 556 (22.7%) | 1216 (20.5%) |
| No | 1922 (77.0%) | 12004 (82.5%) | 1889 (77.3%) | 4727 (79.5%) |
| Missing | 13 (0.5%) | 281 (1.9%) | - | - |
| Pulmonary Disease | | | | |
| Yes | 887 (35.5%) | 4148 (28.5%) | 882 (36.1%) | 1941 (32.7%) |
| No | 1597 (64.0%) | 10120 (69.6%) | 1563 (63.9%) | 4002 (67.3%) |
| Missing | 13 (0.5%) | 281 (1.9%) | - | - |
| Renal Disease | | | | |
| Yes | 349 (14.0%) | 1108 (7.6%) | 343 (14.0%) | 563 (9.5%) |
| No | 2135 (85.5%) | 13160 (90.5%) | 2102 (86.0%) | 5380 (90.5%) |
| Missing | 13 (0.5%) | 281 (1.9%) | - | - |
| Hypertension | | | | |
| Yes | 1305 (52.3%) | 5635 (38.7%) | 1287 (52.6%) | 2720 (45.8%) |
| No | 1179 (47.2%) | 8633 (59.3%) | 1158 (47.4%) | 3223 (54.2%) |
| Missing | 13 (0.5%) | 281 (1.9%) | - | - |
| Obesity | | | | |
| Yes | 814 (32.6%) | 5089 (35.0%) | 808 (33.0%) | 2015 (33.9%) |
| No | 1670 (66.9%) | 9179 (63.1%) | 1637 (67.0%) | 3928 (66.1%) |
| Missing | 13 (0.5%) | 281 (1.9%) | - | - |
| Vaccination Status | | | | |
| Not known to be vaccinated | 1494 (59.8%) | 11524 (79.2%) | 1461 (59.8%) | 3926 (66.1%) |
| Partially vaccinated | 144 (5.8%) | 740 (5.1%) | 140 (5.7%) | 405 (6.8%) |
| Fully vaccinated | 859 (34.4%) | 2285 (15.7%) | 844 (34.5%) | 1612 (27.1%) |
| Pandemic Phase | | | | |
| Pre-Alpha: Nov 2020 - Feb 2021 | 376 (15.1%) | 5991 (41.2%) | 369 (15.1%) | 954 (16.1%) |
| Alpha: March 2021 - June 2021 | 570 (22.8%) | 2764 (19.0%) | 565 (23.1%) | 1494 (25.1%) |
| Delta: July 2021 - Sep 2021 | 1551 (62.1%) | 5794 (39.8%) | 1511 (61.8%) | 3495 (58.8%) |
| Type of Monoclonal Antibody | | | | |
| Bamlanivimab | 401 (16.1%) | - | 394 (16.1%) | - |
| Bamlanivimab + etesevimab | 91 (3.6%) | - | 85 (3.5%) | - |
| Casirivimab + imdevimab | 1987 (79.6%) | - | 1948 (79.7%) | - |
| Sotrovimab | 18 (0.7%) | - | 18 (0.7%) | - |

Abbreviations: mAb, monoclonal antibody; SD, standard deviation

**Appendix Table 5. Primary and Secondary Outcomes by Monoclonal Antibody Treatment Status for Sensitivity Analysis Cohort 1**

| Outcome | mAb-Treated | mAb-Untreated | Adjusted OR | 95% CI |
|---|---|---|---|---|
| **Overall Sample Size** | **n=2445** | **n=5943** | | |
| All-Cause Hospitalization | | | | |
| 28-day | 106 (4.3) | 482 (8.1) | 0.47 | (0.38, 0.59) |
| 90-day | 136 (5.6) | 544 (9.2) | 0.54 | (0.44, 0.66) |
| All-Cause Mortality | | | | |
| 28-day | 3 (0.1) | 57 (1) | 0.12 | (0.03, 0.32) |
| 90-day | 6 (0.2) | 74 (1.2) | 0.18 | (0.07, 0.39) |
| All ED Visits | | | | |
| 28-day | 482 (19.7) | 1055 (17.8) | 1.19 | (1.05, 1.36) |
| **Hospitalized sample size** | **n=106** | **n=482** | | |
| Hospital LOS in days, mean (SD)* | 5.7 (6.5) | 8.5 (10.1) | 0.67 | (0.53, 0.85) |
| IMV or Death | 5 (4.7) | 87 (18) | 0.2 | (0.07, 0.47) |
| ICU Admission | 13 (12.3) | 98 (20.3) | 0.55 | (0.27, 1.02) |
| ICU LOS in days, mean (SD)* | 3.5 (2.8) | 8.6 (9.9) | 0.24 | (0.11, 0.52) |

* Poisson regressions presented as adjusted incidence rate ratios with 95% confidence intervals

All regressions adjusted for age, sex, race/ethnicity, BMI, immunocompromised status, number of other comorbidities, insurance status, pandemic phase, and vaccination status

Abbreviations: mAb, monoclonal antibody; OR, odds ratio; CI, confidence interval; LOS, length of stay; ICU, intensive care unit; SD, standard deviation

**Appendix Table 6. Baseline Characteristics by Monoclonal Antibody Treatment Status for Sensitivity Analysis Cohort 2**

| | Full Cohort | | Matched Cohort | |
|---|---|---|---|---|
| | mAb-Treated (n=2896) | mAb-Untreated (n=33319) | mAb-Treated (n=2797) | mAb-Untreated (n=6864) |
| Age in years* | | | | |
| 18-54 | 1107 (38.2%) | 25075 (75.3%) | 1068 (38.2%) | 3194 (46.5%) |
| 55-64 | 611 (21.1%) | 4669 (14.0%) | 592 (21.2%) | 1647 (24.0%) |
| ≥65 | 1178 (40.7%) | 3575 (10.7%) | 1137 (40.7%) | 2023 (29.5%) |
| Sex* | | | | |
| Female | 1571 (54.2%) | 17681 (53.1%) | 1525 (54.5%) | 3762 (54.8%) |
| Male | 1325 (45.8%) | 15636 (46.9%) | 1272 (45.5%) | 3102 (45.2%) |
| Missing | 0 (0.0%) | 2 (0.0%) | - | - |
| Race/Ethnicity* | | | | |
| Non-Hispanic White | 2341 (80.8%) | 22311 (67.0%) | 2322 (83.0%) | 5481 (79.9%) |
| Hispanic | 275 (9.5%) | 5263 (15.8%) | 272 (9.7%) | 811 (11.8%) |
| Non-Hispanic Black | 65 (2.2%) | 1488 (4.5%) | 65 (2.3%) | 192 (2.8%) |
| Other | 139 (4.8%) | 2326 (7.0%) | 138 (4.9%) | 380 (5.5%) |
| Missing | 76 (2.6%) | 1931 (5.8%) | - | - |
| Insurance Status* | | | | |
| Medicaid | 173 (6.0%) | 3601 (10.8%) | 1421 (50.8%) | 3997 (58.2%) |
| Medicare | 1129 (39.0%) | 3540 (10.6%) | 1096 (39.2%) | 1992 (29.0%) |
| None/Uninsured | 64 (2.2%) | 1728 (5.2%) | 170 (6.1%) | 537 (7.8%) |
| Other/Unknown | 64 (2.2%) | 2142 (6.4%) | 46 (1.6%) | 140 (2.0%) |
| Private/Commercial | 1466 (50.6%) | 22308 (67.0%) | 64 (2.3%) | 198 (2.9%) |
| Body Mass Index, kg/m$^2$ * | | | | |
| <18.5 | 23 (0.8%) | 204 (0.6%) | 23 (0.8%) | 58 (0.8%) |
| 18.5-24.9 | 374 (12.9%) | 3784 (11.4%) | 371 (13.3%) | 856 (12.5%) |
| 25.0-29.9 | 600 (20.7%) | 4666 (14.0%) | 593 (21.2%) | 1398 (20.4%) |
| ≥30.0 | 808 (27.9%) | 5668 (17.0%) | 794 (28.4%) | 2028 (29.5%) |
| Missing | 1091 (37.7%) | 18997 (57.0%) | 1016 (36.3%) | 2524 (36.8%) |
| Immunocompromised* | | | | |
| Yes | 847 (29.2%) | 3281 (9.8%) | 832 (29.7%) | 1669 (24.3%) |
| No | 2026 (70.0%) | 27993 (84.0%) | 1965 (70.3%) | 5195 (75.7%) |
| Missing | 23 (0.8%) | 2045 (6.1%) | - | - |
| Number of Other Comorbid Conditions* | | | | |
| 0 | 807 (27.9%) | 18488 (55.5%) | 759 (27.1%) | 1980 (28.8%) |
| 1 | 733 (25.3%) | 6887 (20.7%) | 723 (25.8%) | 1997 (29.1%) |
| ≥2 | 1332 (46.0%) | 5890 (17.7%) | 1315 (47.0%) | 2887 (42.1%) |
| Missing | 24 (0.8%) | 2054 (6.2%) | - | - |
| Diabetes | | | | |
| Yes | 588 (20.3%) | 2523 (7.6%) | 578 (20.7%) | 1154 (16.8%) |
| No | 2284 (78.9%) | 28742 (86.3%) | 2219 (79.3%) | 5710 (83.2%) |
| Missing | 24 (0.8%) | 2054 (6.2%) | - | - |
| Cardiovascular Disease | | | | |

| | | | | |
|---|---|---|---|---|
| Yes | 583 (20.1%) | 2346 (7.0%) | 574 (20.5%) | 1286 (18.7%) |
| No | 2289 (79.0%) | 28919 (86.8%) | 2223 (79.5%) | 5578 (81.3%) |
| Missing | 24 (0.8%) | 2054 (6.2%) | - | - |
| Pulmonary Disease | | | | |
| Yes | 924 (31.9%) | 5628 (16.9%) | 915 (32.7%) | 2137 (31.1%) |
| No | 1948 (67.3%) | 25637 (76.9%) | 1882 (67.3%) | 4727 (68.9%) |
| Missing | 24 (0.8%) | 2054 (6.2%) | - | - |
| Renal Disease | | | | |
| Yes | 356 (12.3%) | 1108 (3.3%) | 350 (12.5%) | 604 (8.8%) |
| No | 2516 (86.9%) | 30157 (90.5%) | 2447 (87.5%) | 6260 (91.2%) |
| Missing | 24 (0.8%) | 2054 (6.2%) | - | - |
| Hypertension | | | | |
| Yes | 1357 (46.9%) | 6321 (19.0%) | 1336 (47.8%) | 2904 (42.3%) |
| No | 1515 (52.3%) | 24944 (74.9%) | 1461 (52.2%) | 3960 (57.7%) |
| Missing | 24 (0.8%) | 2054 (6.2%) | - | - |
| Obesity | | | | |
| Yes | 837 (28.9%) | 5089 (15.3%) | 829 (29.6%) | 2060 (30.0%) |
| No | 2035 (70.3%) | 26176 (78.6%) | 1968 (70.4%) | 4804 (70.0%) |
| Missing | 24 (0.8%) | 2054 (6.2%) | - | - |
| Vaccination Status | | | | |
| Not known to be vaccinated | 1777 (61.4%) | 28250 (84.8%) | 1707 (61.0%) | 4477 (65.2%) |
| Partially vaccinated | 147 (5.1%) | 1630 (4.9%) | 141 (5.0%) | 460 (6.7%) |
| Fully vaccinated | 972 (33.6%) | 3439 (10.3%) | 949 (33.9%) | 1927 (28.1%) |
| Pandemic Phase | | | | |
| Pre-Alpha: Nov 2020 - Feb 2021 | 402 (13.9%) | 16926 (50.8%) | 390 (13.9%) | 997 (14.5%) |
| Alpha: March 2021 - June 2021 | 642 (22.2%) | 7995 (24.0%) | 630 (22.5%) | 1733 (25.2%) |
| Delta: July 2021 - Sep 2021 | 1852 (64.0%) | 8398 (25.2%) | 1777 (63.5%) | 4134 (60.2%) |
| Type of Monoclonal Antibody | | | | |
| Bamlanivimab | 425 (14.7%) | - | 413 (14.8%) | - |
| Bamlanivimab + etesevimab | 98 (3.4%) | - | 90 (3.2%) | - |
| Casirivimab + imdevimab | 2344 (80.9%) | - | 2265 (81.0%) | - |
| Sotrovimab | 29 (1.0%) | - | 29 (1.0%) | - |

* *Variables* used in the propensity matching. Abbreviations: mAb, monoclonal antibody, SD, standard deviation

**Appendix Table 7. Primary and Secondary Outcomes by Monoclonal Antibody Treatment Status for Sensitivity Analysis Cohort 2**

| Outcome | mAb-Treated | mAb-Untreated | Adjusted OR | 95% CI |
|---|---|---|---|---|
| **Overall Sample Size** | **n=2797** | **n=6864** | | |
| All-Cause Hospitalization | | | | |
| 28-day | 103 (3.7) | 508 (7.4) | 0.45 | (0.36, 0.56) |
| 90-day | 138 (4.9) | 578 (8.4) | 0.53 | (0.43, 0.64) |
| All-Cause Mortality | | | | |
| 28-day | 2 (0.1) | 61 (0.9) | 0.07 | (0.01, 0.24) |
| 90-day | 6 (0.2) | 83 (1.2) | 0.17 | (0.07, 0.36) |
| All ED Visits | | | | |
| 28-day | 527 (18.8) | 1121 (16.3) | 1.28 | (1.13, 1.45) |
| **Hospitalized sample size** | **103** | **508** | | |
| Hospital LOS in days, mean (SD)* | 5.9 (6.6) | 8.4 (10) | 0.68 | (0.54, 0.87) |
| IMV or Death | 5 (4.9) | 89 (17.5) | 0.21 | (0.07, 0.50) |
| ICU Admission | 13 (12.6) | 104 (20.5) | 0.52 | (0.26, 0.96) |
| ICU LOS in days, mean (SD)* | 3.5 (2.8) | 8.6 (9.8) | 0.23 | (0.11, 0.48) |

* Poisson regressions presented as adjusted incidence rate ratios with 95% confidence intervals

All regressions adjusted for age, sex, race/ethnicity, BMI, immunocompromised status, number of other comorbidities, insurance status, pandemic phase, and vaccination status

Abbreviations: mAb, monoclonal antibody; OR, odds ratio; CI, confidence interval; LOS, length of stay; ICU, intensive care unit; SD, standard deviation

**Appendix Table 8. Monoclonal Antibody Emergency Use Authorization Eligibility Criteria**

| Prior to June 1, 2021 | After June 1, 2021 |
|---|---|
| Body mass index of 35 kg/$m^2$ or more | Body mass index of 25 kg/$m^2$ or more |
| Chronic kidney disease | Chronic kidney disease |
| Diabetes | Diabetes |
| Immunosuppressive disease or currently receiving immunosuppressive treatment | Immunosuppressive disease or are currently receiving immunosuppressive treatment |
| 65 years of age or older | 65 years of age or older |
| 55 years of age or older AND have either cardiovascular disease OR hypertension OR chronic obstructive pulmonary disease/other chronic respiratory disease | Chronic respiratory diseases, cardiovascular disease, or hypertension |
| | Pregnancy |
| | Sickle cell disease |
| | Neurodevelopmental disorders |
| | Medical related technology dependence |
| | Other medical conditions or factors (eg, race or ethnicity) that places individual patients at risk for progression to severe COVID-19 |
